# Long stability of SARS-CoV-2 RNA in dry and saliva swab samples stored for diagnostics, Denmark

**DOI:** 10.1101/2021.02.11.21251563

**Authors:** Alonzo Alfaro-Núñez, Stephanie Crone, Shila Mortensen, Maiken Worsøe Rosenstierne, Anders Fomsgaard, Ellinor Marving, Sofie Holdflod Nielsen, Michelle Grace Pinto Jørgensen, Arieh S. Cohen, Claus Nielsen

**Affiliations:** Department of Virus and Microbiological Special Diagnostics, Statens Serum Institut, Artillerivej 5, 2300 Copenhagen S, Denmark; SSI Diagnostica, Herredsvejen 2, 3400 Hillerød, Denmark; TestCenter Danmark, Statens Serum Institut, Artillerivej 5, 2300 Copenhagen S, Denmark

**Keywords:** Viral survival, COVID-19, surveillance, clinical samples, oropharyngeal, saliva, RNA quantification methods

## Abstract

During the current COVID-19 pandemic, Danish authorities demand that clinical swab samples must be analysed within 72 hours. This time constraint was adopted to guarantee a quick and efficient response. As today, analysis results are given within 20 hours average fulfilling this demand. However, it is unclear for how long and under which conditions swab specimens can be stored without clinically affecting SARS-CoV-2 RNA stability. This study demonstrates that swabs specimens can be stored at −20°C and +4°C for up to 26 days without affecting RT-qPCR results. However, early detection of asymptomatic cases is important to reduce spreading COVID-19 may be the final consequence not only in Denmark but worldwide.

## Rapid increase of SARS-CoV-2 clinical sampling

As the number of cases of COVID-19, severe acute respiratory syndrome Coronavirus 2 (SARS-CoV-2), keep rapidly increasing during the current pandemic, there are no signs of the disease slowing down during a the second or even third wave of infection. As such, numbers of clinical samples for detection of SARS-CoV-2 continue to grow worldwide (https://www.ecdc.europa.eu/en/covid-19/data and https://www.cdc.gov/coronavirus/2019-ncov/index.html). The increasing number of samples put pressure on logistics at the various test laboratories and challenges their ability to analyse samples in a timely fashion. While new, innovative and faster methods for screening and diagnosing patients are constantly being developed (e.g. CRISPR-Cas based, and other isothermal amplification methods, conventional antigen-based test for viral proteins, PCR assays for detecting viral amplicons), extraction of nuclei acids, followed by either reverse transcription polymerase chain-reaction (RT-PCR) or real-time RT-PCR (RT-qPCR) approaches, remain the most commonly used due to their high sensitivity and relatively low cost (1).

Healthcare institutions, research labs and governments have made great efforts to maintain the flow of analytical diagnostics on time to provide results for the adequate detection of positive SARS-CoV-2 clinical cases worldwide.

In Denmark, there are two different test tracks available. Hospitals across the country run the health test track and are responsible for testing people with clear COVID-19 symptoms. While Test Centre Denmark (TCDK) at the Statens Serum Institut (SSI) runs the society test track (STT), which allows asymptomatic persons, or people with only mild symptoms, or even people who just wants to know their health status to be tested. One of the main premises while establishing TCDK during the early 2020 was that machines, consumables and reagents used at this facility, must not be the same as the ones used at the health track, to avoid the shortage experienced during the early stages of the pandemic across all Europe, and worldwide. Up today, TCDK runs in average more than 100,000 COVID-19 tests a day as the government recommends every person going physically to work to be tested at least once a week.

Additionally, the established procedure by SSI requires and demands that all COVID-19 clinical swabs have to be analysed within 72 hours after sample collection. In fact, over 90% of the samples are analysed and results are given within 20 hours average. Any sample received and analysed after this time limit is reported as inconclusive and then discarded. The rationale for the 72-hour deadline is based on two principals. The first principal is that for a testing strategy to be effective results must be available as soon as possible. As the final results represent a moment in time, a delayed outcome may no longer signify the actual health status of a patient. Moreover, persons with even slight symptoms are expected to self-quarantine while waiting for the results and this is a substantial burden. The second principal is that when large scale testing was started, it was unknown how long SARS-CoV-2 RNA remains stable on clinical sample swabs. A stability below 72 hours was considered reasonable (2). Furthermore, the laboratories for the STT are now centralized within two facilities across the country, samples are received from all over the nation and may be delayed in transport, which will subsequently cause them to be excluded from screening diagnostic if arrive outside this time threshold. This may represent a loss of potential COVID-19 positive cases carrying any of the different SARS-CoV-2 variants (3), and raises the question to how long does SARS-CoV-2 RNA remain stable in the clinical swab samples during transportation and before arrival to the respective analytical centre?

Despite the large volume of documentation generated in 2020 about the pathogenicity, infectivity and detection of SARS-CoV-2 and COVID-19, there is, however, only a handful of studies evaluating the longevity and conditions for SARS-CoV-2 RNA particle stability maintenance in clinical swabs samples for transportation in different medias (4-6). Furthermore, to our knowledge, there is no available data on the stability of SARS-CoV-2 RNA during transportation and longevity in dry swabs (without any preservation buffer). Therefore, in our current study, we add to the previous findings on viral survival and detection in order to assess the stability at different environmental temperatures of SARS-CoV-2 RNA particles in swab material without the addition of a preservation media.

## Experimental design for viral quantification

A total of 120 swabs (CLASSIQSwabs™ Dry Swabs, COPAN) were spiked with SARS-CoV-2 in the lab under two main conditions, 60 swabs in “Dry” by adding 5 µL of the virus directly to the tip of the swab, and 60 oropharyngeal swabs in “Saliva” solution sampled from one unique person, and then, by also adding 5 µL of SARS-CoV-2. No transport or stabilizing media was added in either of the two conditions, thus, both should be considered dry swabs. Swab samples were stored at three different temperatures (−20°C, +4°C and +20°C), and thereafter analysed in triplicate by RT-qPCR after 1, 3, 5, 8, 9, 15 and 26 days. All incubations took place in dark to overrule the potential effect of light UV radiation in the degradation of RNA molecules.

A well-characterized strain of SARS-CoV-2 (2019-nCoV/Munchen 1-2-2020/964, Charite/Berlin) diluted in PBS, was used to spike the swabs with a *C*_T_ -value of 29,4 (∼10^3^ viral copies/ µL). Dilution series of the original standard sample (*C*_T_ -value of 20; ∼10^6^ viral copies/ µL) allowed the estimation of RNA viral copies/µL.

Samples were analysed at TCDK where 700 µL of PBS were added to the swabs, and the samples were left agitating on a shaker for 10 min (700RPM). Total nucleic acids were extracted from 200 µL of sample using a Biomek i7 (Beckman Coulter) and the RNAdvance Blood kit (Beckman Coulter) following the manufacturer guidelines and eluting in 50 µL DNase and RNase free water. RT-qPCR was performed on a CFX96 (Bio-Rad) using 5 µL elute in a total reaction volume of 25 µL. The PCR reaction contains Luna Probe One-Step React Mix (New England Biolabs), 0,3% IGEPAL CA-630. The Primers & Probes targeting the E-gene region and Cycling conditions as described in Corman *et al*. 2020 (7). This standard set up for analysing all the SST swab samples keeps being updated and improved day by day.

## How long does SARS-CoV-2 survive in oropharyngeal dry swab clinical samples?

The design of our study differed from previous investigations that compared swab types and specimen collection methods within a clinical setting (4,8). Despite the fact that nasopharyngeal swabs are considered to reach the highest-yield for diagnostic testing of SARS-CoV-2, oropharyngeal swabs and saliva are also broadly used (9). Studies suggest that saliva may be a suitable and high-yield diagnostic type of sample for detection of SARS-CoV-2. According to local viral replication, in addition to the potential mixing in saliva of lower and upper respiratory tract fluids that can carry virus (9,10). Our approach was laboratory based and no patients were involved. The analysis was only designed as a development study, using the current oropharyngeal national swab method, to provide insight into the stability of RNA viral particles in dry and saliva swabs and not in the disease itself. Even though *C*_T_ values provide quantitative information over time, issues such as the relationship between viral load and disease severity could therefore not be assessed and were never the target of the current assay.

In overall, no significant variation was observed in the *C*_T_ values (and viral copy number/ µL) over time within the two main conditions (see Fig. 1), except for the saliva treatment at +20°C which after 9 days experienced an increase in the mean *C*_T_ values (from ∼10^3^ to ∼10^1^ viral copies / µL concentration decrease). A direct correlation of higher viral stability in lower temperatures over time for both main conditions was observed (Table 1). Moreover, there was also a slight tendency of saliva swab samples to give lower *C*_T_ values and thus, to be a more stable environment for the survival of SARS-CoV-2 RNA during the first 9 days of incubation. A study showed that SARS-CoV-2 could be detected using RT-qPCR on swabs after 21 days at room temperature (8). In another study, absolute dry swabs taken from clinical patients showed a slight reduction in utility for several respiratory viruses, supporting our results (2). Although, both conditions in our study should be essentially considered as dry swabs as no preservation buffer were added. The saliva swabs represented better the actual enzymatic environment found in real clinical swab samples, than the absolute dry swabs, in which only 5 µL of the virus was added directly to the tip of the swab. Nevertheless, to our knowledge there is no study evaluating the direct effect of oropharyngeal enzymes in the stability of SARS-CoV-2 and thus, we cannot further conclude.

**Table 1.** Statistical comparison of *C*_T_ values per conditions. *C*_T_ mean value results with the SD (standard deviation) values for the two main conditions, in” saliva” medium and in dry (no medium). Swabs were spiked with 5 µL of SARS-CoV-2 cultivated virus and quantified after 1, 3, 5, 8, 9, 15 and 26 days. Simultaneously, for each treatment, three different environmental temperature were evaluated (−20°C, 4°C and +20°C). Moreover, three replicas were quantified within each treatment in addition to a negative control. Finally, no transport or stabilizing media was added to neither of the two conditions, thus, both should be considered dry swabs.

| SARS-CoV-2 in saliva medium |  |  |  |  |  |  |  |
| --- | --- | --- | --- | --- | --- | --- | --- |
|  | Mean CT (SD) on day: |  |  |  |  |  |  |
|  | 1 | 3 | 5 | 8 | 9 | 15 | 26 |
| -20°C | 31,4 (0,7) | 32,3 (1,1) | 30,8 (0,4) | 32,2 (0,9) | 31,6 (1,4) | 31,8 (0,4) | 31,5 (0,5) |
| 4°C | 30,9 (1,3) | 31,1 (0,3) | 30,9 (1,3) | 31,0 (0,1) | 31,2 (0,3) | 32,0 (0,6) | 32,0 (0,06) |
| 20°C | 30,8 (0,5) | 29,5 (1,7) | 31,8 (2,1) | 30,7 (0,7) | 31,3 (0,8) | 34,6 (0,3) | 33,4 (1,0) |
|  | P values day: |  |  |  |  |  |  |
|  | 1 | 3 | 5 | 8 | 9 | 15 | 26 |
| -20°C vs. 4°C | 0,76 | 0,32 | 1 | 0,38 | 0,91 | 0,96 | 0,79 |
| -20°C vs. 20°C | 0,73 | 0,003 | 0,43 | 0,14 | 0,93 | 0,003 | 0,05 |
| 4°C vs. 20°C | 1,00 | 0,11 | 0,49 | 0,92 | 1,00 | 0,006 | 0,21 |
| P values day 1 vs day 9 |  |  |  | P values day 1 vs day 26 |  |  |  |
| -20°C | 1,00 |  |  | 1,00 |  |  |  |
| 4°C | 1,00 |  |  | 0,73 |  |  |  |
| 20°C | 1,00 |  |  | 0,03 |  |  |  |

| SARS-CoV-2 no medium dry swab |  |  |  |  |  |  |  |
| --- | --- | --- | --- | --- | --- | --- | --- |
|  | Mean CT (SD) on day: |  |  |  |  |  |  |
|  | 1 | 3 | 5 | 8 | 9 | 15 | 26 |
| -20°C | 31,8 (0,6) | 31,9 (0,8) | 31,8 (2,0) | 31,1 (0,4) | 31,8 (0,48) | 31,2 (1,4) | 31,9 (0,3) |
| 4°C | 33,3 (0,4) | 32,8 (2,0) | 31,6 (0,4) | 33,2 (0,4) | 32,7 (0,5) | 31,2 (1,6) | 32,1 (1,3) |
| 20°C | 32,4 (0,8) | 33,8 (0,3) | 32,7 (1,3) | 34,0 (1,7) | 33,1 (0,7) | 32,5 (0,5) | 33,2 (0,6) |
|  | P values day: |  |  |  |  |  |  |
|  | 1 | 3 | 5 | 8 | 9 | 15 | 26 |
| -20°C vs. 4°C | 0,21 | 0,51 | 0,96 | 0,04 | 0,63 | 1,00 | 0,99 |
| -20°C vs. 20°C | 0,77 | 0,08 | 0,54 | 0,004 | 0,34 | 0,33 | 0,32 |
| 4°C vs. 20°C | 0,55 | 0,51 | 0,38 | 0,67 | 0,85 | 0,32 | 0,41 |
| P values day 1 vs day 9 |  |  |  | P values day 1 vs day 26 |  |  |  |
| -20°C | 1,00 |  |  | 1,00 |  |  |  |
| 4°C | 0,99 |  |  | 0,77 |  |  |  |
| 20°C | 0,98 |  |  | 0,97 |  |  |  |

**Figure 1.**
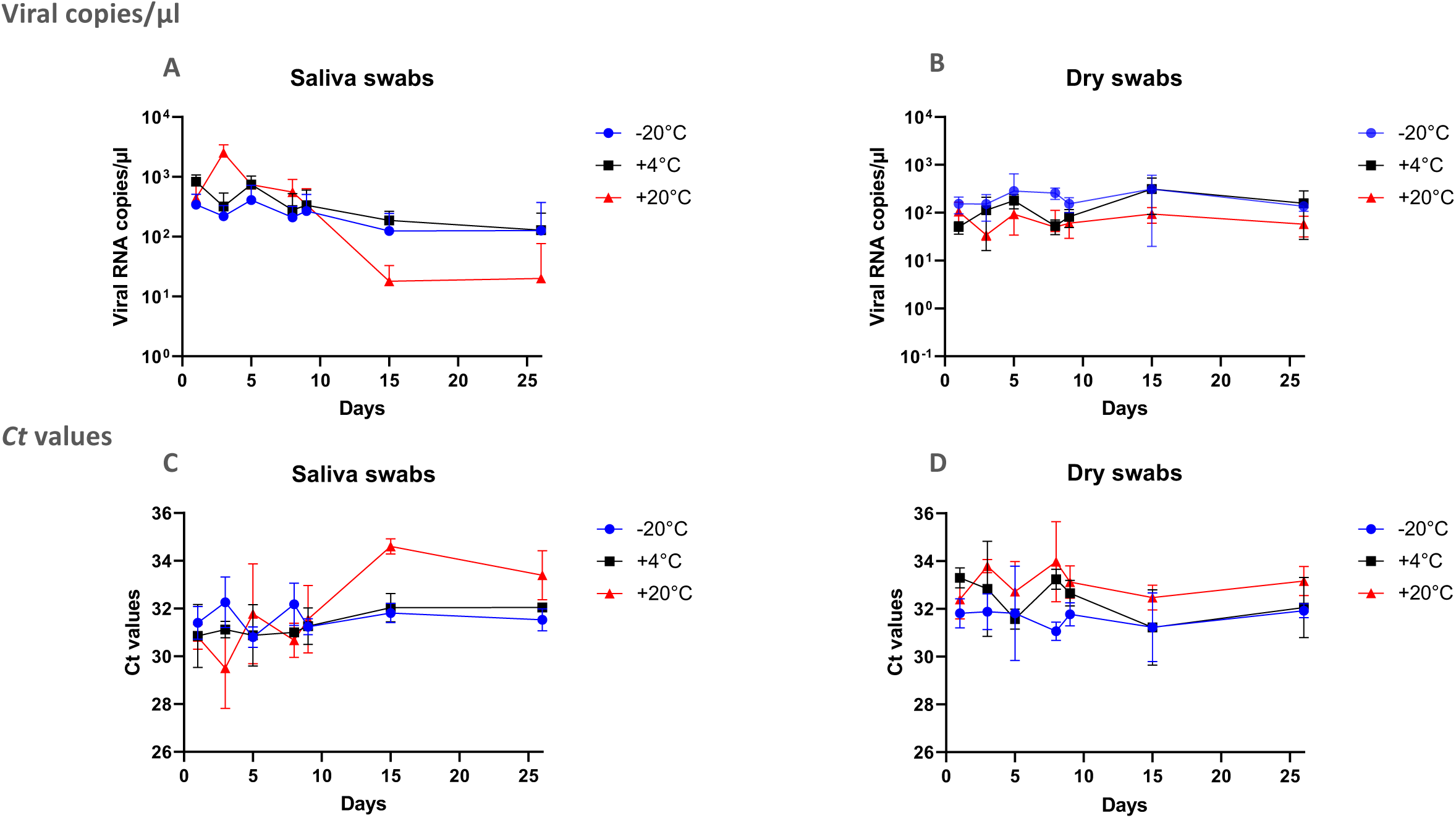
*C*_T_ values variation and RNA viral copies / µL concentrations in dry and saliva swabs over time. RNA viral copies / µL mean concentrations over time are presented in panels **A** for saliva swabs and panel **B** for dry swabs. *C*_T_ value mean results for the two main categories, in panel **C** for saliva medium and panel **D** for absolutely dry. Swabs were spiked with 5uL of SARS-CoV-2 cultivated virus and quantified after 1, 3, 5, 8, 9, 15 and 26 days. Simultaneously and for each treatment, three different environmental temperature were evaluated at −20°C, 4°C and +20°C. Three (3) replicas were quantified within each treatment in addition to a negative control. No transport or stabilizing media was added to neither of the two conditions, thus, both should be considered dry swabs.

As mentioned above, temperature was observed to play a significant role in stabilizing the RNA molecules of the virus, in particular for the saliva swabs when compared to the dry swabs (Suppl. Mat. 1). Using linear regression, there was a statistically significant difference in *C*_T_ in the saliva swabs after 3 days (*P* = 0,003), 15 days (*P* = 0,003) and 26 days (*P* = 0,03) of incubation, and in dry swabs after 8 days (*P* = 0,004) incubation in between −20°C and ambient room temperature (20°C). Moreover, a significant difference was also noticed for the saliva after 15 days (*P* = 0,006) when compared the +4°C and +20°C temperatures, and in the dry treatment after 8 days (*P* = 0,03) in between the −20°C and +4°C incubation temperature (Table1). Finally, samples at −20°C were slightly more stable than +4°C and samples at +20°C showed increase in viral particles degradation with lower viral copies (Suppl. Mat. 2).

There are several available protocols and regulations for oropharyngeal swab samples collection and the different media available for transportation (11,12). Oropharyngeal swabs are among the most used type of clinical sample specimens collected in combination with nasopharyngeal swabs during the current pandemic (6). In Denmark, hospitals with the health track use different kinds of transport buffer containing a hydrolysing agent to the swabs right after collection. However, SSI with the STT does not add a transport buffer and swab samples are stored dry after collection until they reach TCDK for nuclei acid extraction and RT-qPCR analysis. There is evidence supporting that transporting dry swabs do not compromise RNA recovery from clinical samples (2). Our results confirm that the current method selected for the STT do not compromise and retain SARS-CoV-2 RNA up to 26 days (a highly surprising number of days) without a significant variation or reduction in *C*_T_-values if the samples are kept cold. As such, implementing the use of dry swabs represents also an economical value by reducing the cost of additional preservation buffers. Furthermore, the swabs used by TCDK were selected based on a preliminary assay that quantified the retention of viral particles by comparing different types of swab materials (Suppl. Mat. 3).

Denmark has become a worldwide reference for developing performance in detection of COVID-19 (8). Testing delays and laboratory bottlenecks have contributed to the aggravation spread of COVID-19 infections. Therefore, an important aspect to take into consideration supporting the innovative implementation of STT is the reduction of spreading COVID-19 by the early detection of asymptomatic cases.

As such, to have a good knowledge of stability survival of SARS-CoV-2 RNA in dry swabs can serve as reference guideline and important for planning during the current pandemic and future events.

This study demonstrates that swabs specimens can be stored at −20°C and +4°C for up to 26 days without clinically affecting RT-qPCR results.

## Supporting information

Suppl. Mat. 1

Suppl. Mat. 2

Suppl. Mat. 3

## Data Availability

All data generated in this study is provided as supplemental materials.

## Ethical statement

Exemption for review by the ethical committee system and informed consent was given by the Committee on Biomedical Research Ethics - Capital region in accordance with Danish law on assay development projects (see Journal-nr.: H-21000338).

## Acknowledgements

We would like to extend our gratitude to Susanne Lopez Rasmussen and Bettina Andersen for their technical assistance.

## Author contributions

Alonzo Alfaro-Núñez (AAN): conceptualization, writing original draft, analysis and validation, approved final MS

Stephanie Crone (SC): review and edit, statistical analysis, approved final MS Sofie Holdflod Nielsen (SHN), Michelle Jørgensen (MJ): generation of data, approved final MS

Shila Mortensen (SM), Arieh S. Cohen (ASC) and Claus Nielsen (CN): conceptualization, review and edit, approved final MS

Maiken Worsøe Rosenstierne (MWR), Anders Fomsgaard (AF) and Ellinor Marving (EM): review and edit, approved final MS

## Table, Figure and Supplemental materials legends

**Supplemental Material 1**. Statistical analysis of *C*_T_ values.

**Supplemental Material 2**. Standard curve RNA viral copies / µL concentrations.

**Supplemental Material 3**. Detection of SARS-CoV-2 using three commercial swab types quantifying the swab retention performance.

## References

1. D’Cruz RJ, Currier AW, Sampson VB. Laboratory Testing Methods for Novel Severe Acute Respiratory Syndrome-Coronavirus-2 (SARS-CoV-2). Front Cell Dev Biol. Frontiers; 2020; 8.

2. Moore C, Corden S, Sinha J, Jones R. Dry cotton or flocked respiratory swabs as a simple collection technique for the molecular detection of respiratory viruses using real-time NASBA. Journal of Virological Methods. Elsevier; 2008; 153(2):84–89.

3. Spiess K, Alfaro-Núñez A, Marving E, et al. Development of RT-PCR to detect the multigeographical variants in the SARS-CoV-2 spike glycoprotein gene. In preparation.

4. Rogers AA, Baumann RE, Borillo GA, et al. Evaluation of Transport Media and Specimen Transport Conditions for the Detection of SARS-CoV-2 by Use of Real-Time Reverse Transcription-PCR. McAdam AJ, editor. J Clin Microbiol. American Society for Microbiology Journals; 2020; 58(8):1564.

5. Ren S-Y, Wang W-B, Hao Y-G, et al. Stability and infectivity of coronaviruses in inanimate environments. WJCC. 2020; 8(8):1391–1399.

6. Perchetti GA, Huang M-L, Peddu V, Jerome KR, Greninger AL, McAdam AJ. Stability of SARS-CoV-2 in Phosphate-Buffered Saline for Molecular Detection. McAdam AJ, editor. J Clin Microbiol. American Society for Microbiology Journals; 2020; 58(8).

7. Corman VM, Landt O, Kaiser M, et al. Detection of 2019 novel coronavirus (2019-nCoV) by real-time RT-PCR. Eurosurveillance. European Centre for Disease Prevention and Control; 2020; 25(3):2000045.

8. Skalina KA, Goldstein DY, Sulail J, et al. Extended storage of SARS-CoV-2 nasopharyngeal swabs does not negatively impact results of molecular-based testing across three clinical platforms. Journal of Clinical Pathology. BMJ Publishing Group; 2020.

9. Lee RA, Herigon JC, Benedetti A, Pollock NR, Denkinger CM. Performance of Saliva, Oropharyngeal Swabs, and Nasal Swabs for SARS-CoV-2 Molecular Detection: A Systematic Review and Meta-analysis. J Clin Microbiol. 2021.

10. Chen L, Zhao J, Peng J, et al. Detection of SARS□CoV□2 in saliva and characterization of oral symptoms in COVID□19 patients. Cell Prolif. John Wiley & Sons, Ltd; 2020; 53(12):1539.

11. Druce J, Garcia K, Tran T, Papadakis G, Birch C. Evaluation of Swabs, Transport Media, and Specimen Transport Conditions for Optimal Detection of Viruses by PCR. J Clin Microbiol. 5 ed. American Society for Microbiology Journals; 2012; 50(3):1064–1065.

12. Rodino KG, Espy MJ, Buckwalter SP, et al. Evaluation of Saline, Phosphate-Buffered Saline, and Minimum Essential Medium as Potential Alternatives to Viral Transport Media for SARS-CoV-2 Testing. McAdam AJ, editor. J Clin Microbiol. American Society for Microbiology Journals; 2020; 58(6).

