## Supplementary material for "Long stability of SARS-CoV-2 RNA in dry and saliva swab samples stored for diagnostics, Denmark": Suppl. Mat. 3

**Supplemental Material 3**. Detection of SARS-CoV-2 using three commercial swab types quantifying the swab retention performance.

The ability to absorb and release SARS-CoV-2 virus was compared for three swab types: CLASSIQSwab (dry rayon, Copan), FLOQSwab (dry flocked swab, Copan) and ST flocked swab (dry flocked swab, Sun Trine).

SARS-CoV-2 (isolate USA-WA1/2020, heat inactivated, ZeptoMetrix) dilutions were prepared in saline.

Swabs were immersed in 150 µl virus dilution for 10 seconds, transferred to 700 µl PBS and swirled for 10 seconds. For each concentration 10 swabs were tested.

RNA was extracted from 200 µl of sample using PureLink™ Viral RNA/DNA Mini Kit (ThermoFisher Scientific) with an elution volume of 50 µl.

SARS-CoV-2 was quantitatively detected using RT-PCR (Aridia COVID-19 Real-Time PCR Test, CTK) according to the manufacturer’s instruction. The assay targets the viral *N* gene and the *ORF1ab* gene and separate Ct values for the two genes were obtained.

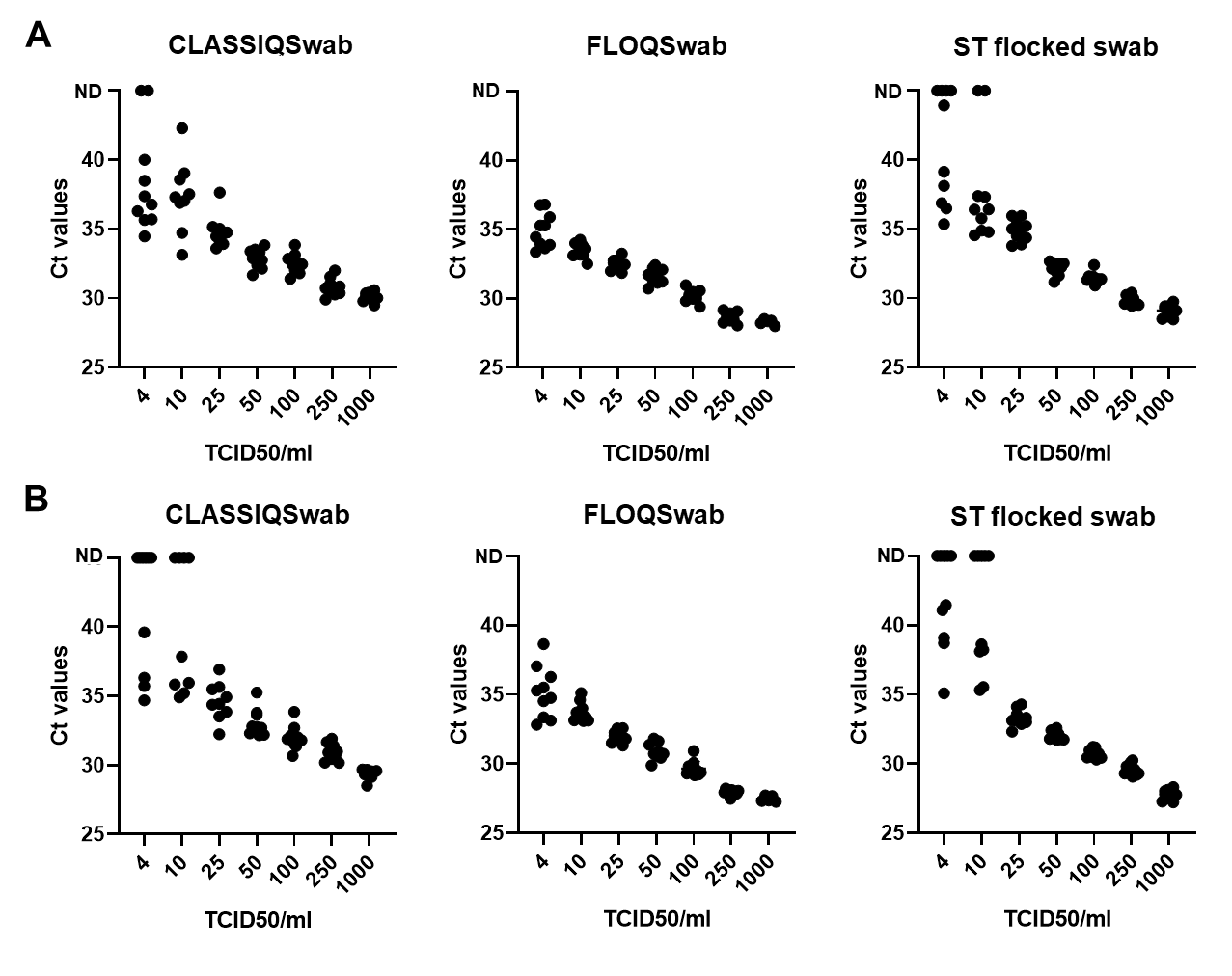

**Figure S1** Ct values for SARS-CoV-2 from three types of swabs inoculated with varying SARS-CoV-2 virus concentrations. (A) *N* gene detection. (B) *ORF1ab* gene detection. ND; not detected.
